# Spatiotemporal modelling of cholera and implications for its control, Uvira, Democratic Republic of the Congo

**DOI:** 10.1101/2023.08.22.23294124

**Authors:** Ruwan Ratnayake, Jacqueline Knee, Oliver Cumming, Jaime Mufitini Saidi, Baron Bashige Rumedeka, Flavio Finger, Andrew S. Azman, W. John Edmunds, Francesco Checchi, Karin Gallandat

## Abstract

The African Great Lakes region including Eastern Democratic Republic of the Congo is a hotspot for cholera transmission. We evaluated the local and global clustering of cholera using 5 years (2016—2020) of suspected cases positive by rapid diagnostic test in Uvira, South Kivu to detect spatiotemporal clusters and the extent of zones of increased risk around cases. We detected 26 clusters (mean radius 652m and mean duration 24.8 days) which recurred annually in three locations and typically preceded seasonal outbreaks. We found a 1100m zone of increased infection risk around cases during the 5 days following clinic attendance for the 2016—2020 period and a 600m radius risk zone for 2020 alone. These risk zone sizes correspond with the area typically used for targeted intervention in the Democratic Republic of the Congo. Our findings underscore the value of the site-specific evaluation of clustering to guide targeted control efforts.

## INTRODUCTION

Cholera outbreaks continue to impact communities that lack access to safe water and adequate sanitation.(1–4) In these communities, cholera’s relatively high reproduction number and short median incubation period (1.4 days, 95% credible interval 1.3—1.6)(1, 5) mean that an initial cluster can rapidly propagate across space. During outbreaks, household fecal-oral transmission through contaminated water, food, and fomites and direct contact becomes substantial and therefore, interventions to prevent infection of household contacts can reduce household transmission.(1, 6–8) Spatiotemporal clustering patterns around affected households have also demonstrated the propagation of transmission between neighboring households.(9, 10) To attenuate and possibly contain transmission during outbreaks based on this natural clustering, case-area targeted interventions (CATI), consisting of an early, multisectoral response within a 100—500m area around case-households, have been proposed.(11–13) CATIs typically include water, sanitation, and hygiene (WASH) interventions to improve water quality and safety (i.e., point of use water treatment, safe drinking water storage containers) and promote hygiene practices like handwashing, antibiotic chemoprophylaxis, and sometimes, oral cholera vaccination (OCV).(13) CATIs with WASH have been a major component of response strategies in Haiti and Yemen(13, 14) while CATIs with WASH and OCV have been used to suppress small outbreaks after mass vaccination campaigns in Juba, South Sudan and Kribi, Cameroon.(15, 16) In the Democratic Republic of the Congo (DRC), similar area-targeted interventions have included the distribution of hygiene kits to case-households and the targeting of WASH interventions to a 500m radius around the most recent cases.(8, 17)

Studies in urban Kalemie, DRC and N’Djamena, Chad have estimated the zone of increased risk of infection around incident cases of at least 200m within the first 5 days after case presentation and in rural Matlab, Bangladesh, up to 450m within the first 3 days after case presentation.(9, 10) As CATIs and other targeted interventions become part of routine public health practice(14), more insight is required into the size and duration of the spatiotemporal zones of increased infection risk required to achieve a substantive impact on transmission, particularly in endemic areas. The size of the zone is likely influenced by factors that determine the strength of community transmission including population density, immunity, vaccination coverage, access to safe water and sanitation, and timeliness of the response.(3, 13)

Cholera has been endemic in the African Great Lakes Region including Eastern DRC since at least 1978 and now contributes substantially to the global cholera burden.(2, 18–22) The *V. cholerae* O1 sublineage AFR10 was introduced from South Asia to East Africa in the late 1990s, and has been driving transmission across the region during the last two decades.(23, 24) In Uvira, South Kivu, DRC, cholera is endemic with stable transmission punctuated by seasonal outbreaks.(18) Using an enhanced clinical surveillance system with rapid diagnostic testing that was setup in Uvira’s cholera treatment units to support an impact evaluation of water supply infrastructure improvements(25), we investigate the location, timing, and annual prediction of spatiotemporal clustering and to estimate the extent of spatiotemporal zones of increased cholera risk around incident cases in an endemic setting.

## METHODS

### Setting

Uvira is a town of approximately 280,000 located on the shore of Lake Tanganyika, an internationally-designated transmission hotspot where suspected cholera cases are reported year-round.(18–20) Seasonal emergence of cholera in Uvira is driven by seasonal exposure to aquatic reservoirs of *V. cholerae* in lakeside waters and person-to-person transmission, excess rainfall linked to fecal contamination of water sources, interruption of water supply and conflict and forced displacement.(18, 20, 26–28) Several city-wide interventions have been implemented including a water supply infrastructure program to improve the production and supply of piped drinking water for which construction started in 2018 and mass vaccination that took place from July to October 2020.(25)

### Data sources

We used a line list of suspected cases (i.e., passing ≥3 loose or watery stools in 24 hours) who received care between 2016 and 2020 at either the Uvira General Hospital’s cholera treatment center (CTC) or the Kalundu cholera treatment unit (CTU, which opened in July 2019). Since April 2016, as part of an evaluation of a water supply infrastructure improvement program, rectal swabs collected from suspected cases have been systematically tested using an RDT (Crystal® VC O1/O139, Arkray Healthcare Pvt. Ltd, Gujarat, India) after a 6 hour enrichment period in alkaline peptone water at ambient temperature.(25, 29) The pooled sensitivity and specificity estimates for enriched Crystal® VC RDTs are 83% (95% CI 67—92) and 98% (95% CI 94—99).(30) We extracted data for 2016—2020, including date of admission, completion of RDT and result, and avenue of residence (i.e., a small census enumeration area of mean size of 1177 (range 180—5711) persons based on population sizes estimated from 2017 official records.(31) The mean avenue area is 0.08 km^2^ and minimum, maximum, and mean distances between avenue centroids are 0.0437m, 12.4km, and 3.1km, respectively. We based the main analyses on enriched RDT-positive cases given the presence of systematic testing in the CTC/CTU and assessments done at the CTC between 2017— 2018 where only 40% of suspected cases were confirmed by polymerase chain reaction.(32)

### Descriptive analysis and seasonal decomposition

We described suspected cases using incidence per 10,000 population, proportion tested with RDT and proportion tested that were RDT-positive. To identify the timing of the cholera season, we analyzed the seasonal decomposition of the weekly incident series of RDT-positive cases with seasonal and trend decomposition using LOESS STL (locally estimated scatterplot smoothing). This method decomposes the time series into trend, seasonal, and random error components based on a two-week trend window and fixed seasonal pattern, and uses the additive model as the seasonal trends appeared relatively constant over time.(33) Missing data were integrated over the case-series with multiple imputation using chain equations.

### Methods for spatiotemporal clustering

We used two different methods to measure spatiotemporal clustering for different phenomena. The *space-time scan statistic* describes local clustering, or the expected density of cases at specific locations within a given area.(34) This gives the timing and locations where cases cluster, exceeding their expected density. The *tau statistic* (τ) describes global clustering, or the overall tendency for cases to occur near other cases in space and time.(35) This suggests the geographic and temporal extents of the zones of increased infection risk. See Appendix Text 1 for the mathematical formulation of these statistics.

### Local clustering to identify recurrent locations and timing of seasonal outbreaks

We used the space-time scan statistic to retrospectively detect the presence and location of spatiotemporal clusters. We conducted the analysis for the entire period (2016—2020) and by year. A relative risk (RR) compares the observed versus expected number of cases inside and outside of a cluster. Poisson distribution of the cases per avenue was assumed. To find the most likely cluster, candidate clusters were ordered by a log-likelihood ratio (LLR) where the cluster with the largest LLR is the least likely to be due to chance and therefore, the most likely cluster. The significance of each cluster was evaluated using Monte Carlo simulation to compare the original dataset with 999 random replicates produced under the null hypothesis.

We examined the entire dataset (i.e., a retrospective scan). We restricted the temporal and spatial windows to capture brief time periods (7—60 days) and a radius that included ≤10% of the population at-risk. To capture clustering that persisted across years, we also used a longer temporal window (7—365 days) for 2016—2020.

To explore whether the space time scan statistic produced signals that preceded outbreaks, we conducted prospective scans of each of the clusters that were detected retrospectively. This was done to detect the earliest warning signal that indicates when that cluster would have first been detected. We simulated repeated prospective scans on the date of the retrospective cluster start day and each successive day (up to 4 weeks later) and calculated the median and IQR of the delay where a prospective scan would have first detected the cluster from the date produced by the retrospective scan that used more case data, and the cluster size at first detection. We visualized on the epidemic curve the timing of the first day of each retrospective cluster. To explore where transmission predominated, we calculated the proportion of the years that the avenue was included in any cluster from 2016—2020, ranging from 0 (not included in any cluster) to 5 (included in a cluster every year).(36)

### Global clustering to inform the boundaries of risk

We estimated the tau statistic (τ) for the entire period (2016—2020) and annually to quantify the spatial extent of the risk zone around an index case.(35) As the dataset only contained the date of the visit to the CTC/CTU as opposed to the date of symptom onset, this represented the risk of developing medically-attended disease, which we assumed indicates severe dehydration/diarrhea (compared with mild dehydration/diarrhea). This approach defines clustering in terms of how likely it is that any pair of cases are potentially transmission-related, within a given distance between the cases. Accordingly, we first classified each pair of cases as potentially transmission-related if their dates of presentation were within 0-4 days of each other (approximately one serial interval).(5) τ is the RR that an individual in the population within a given distance band (d_1_, d_2,_ e.g., 100m, 150m) from an incident case also becomes a case that is potentially transmission-related, compared to the risk of any individual in the population becoming a potentially transmission-related case. τ >1 indicates evidence of clustering within the given distance band.

As we lacked individual household locations of cases, τ reflects the spatial scale of the avenues. We estimated τ with a moving window of 50m computed every 10m at distances from 420m (as 5% of inter-avenue centroids fell below this value) to 2500m (approximate width of Uvira). We calculated the 95% CIs using the 2.5th and 97.5th quantiles from 1000 bootstrap replicates. We evaluated τ over a 5-day window which included the date of case presentation, and a 4-day window which excluded the date of case presentation to account for a more realistic response started the day after.(9) To smooth the artefactual fluctuations resulting from the resolution of the data and smaller sample size of annual datasets, we calculated a moving average over the previous 10m. We defined the high-risk zone around incident cases as the radius up to which the moving average’s lower 95% CIs crossed 1.0 for ≥30 consecutive meters. We defined another elevated-risk zone around incident cases as the radius up to which the moving average point estimate crossed 1.0 for ≥30 consecutive meters. To explore the potential biases from using centroids compared to household locations, we conducted a simulation study where we randomly assigned household locations within each case’s avenue and then estimated τ using a lower distance range (75—2500m) (Appendix Text 2, Figures 1—4).

**Figure 1.**
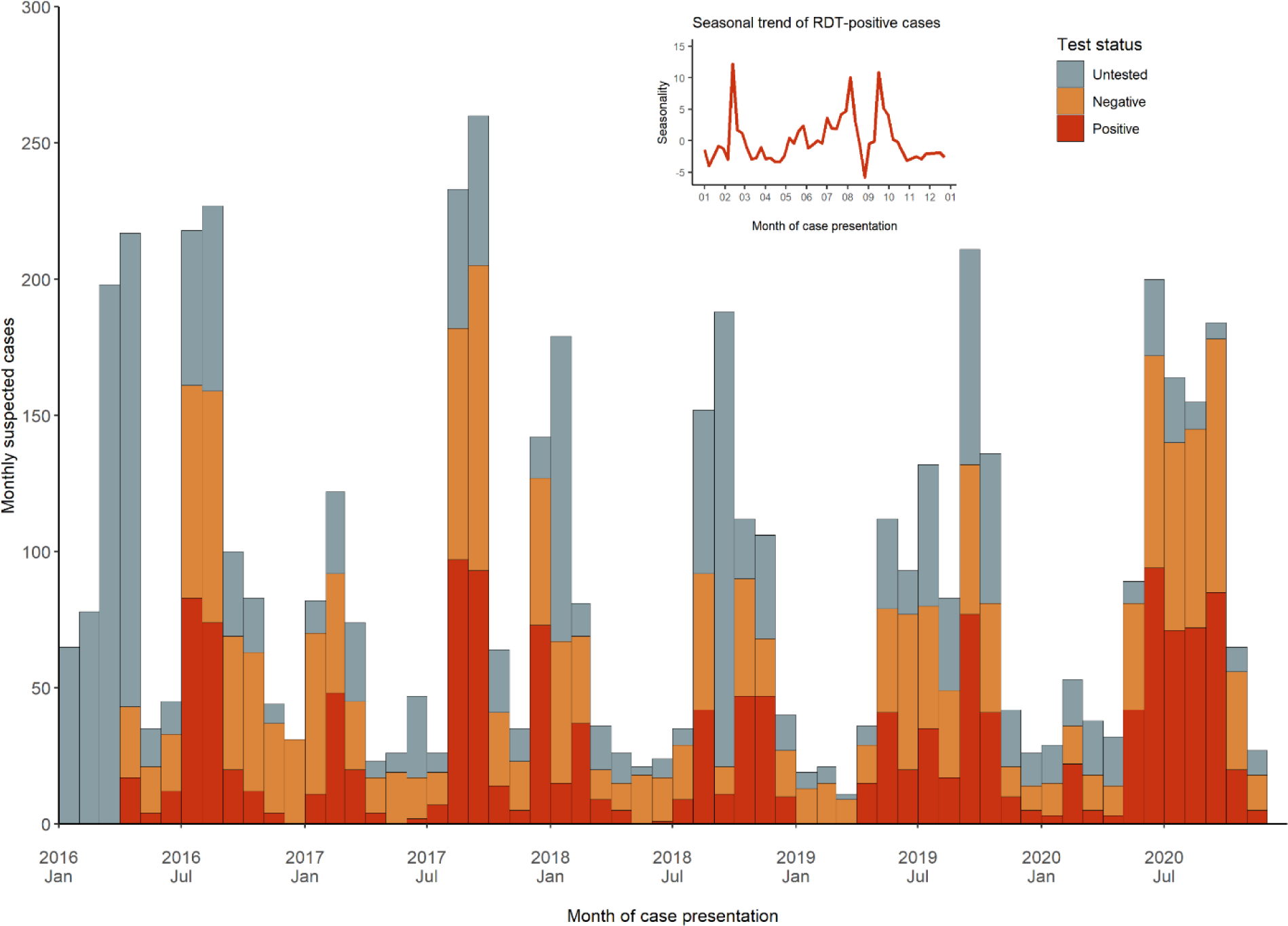
Cholera, Uvira, 2016—2020: Epidemic curve of monthly suspected cases by test status(25) with seasonal trend of monthly RDT-positive cases (inset).

For both the scan statistics and τ, we carried out sensitivity analyses using all suspected cases (i.e., RDT-positive and negative cases, and untested cases that met the suspected case definition).

All analyses were carried out in R software (v. 4.1.2) using the rsatscan (v. 1.0.5) and IDSpatialStats (v. 0.3.12) R packages to calculate the scan statistic (with SaTScan™ v. 10.0.2 software) and τ.(37–40) Ethical approval was provided by the London School of Hygiene and Tropical Medicine (#10603-5) and the University of Kinshasa School of Public Health (#ESP/CE/173B/2022).

## RESULTS

5,447 suspected cases were recorded from 2016 to 2020. 3,456 (63.4%) of the 5,447 suspected cases were tested, of which 1,493 (43.2%) of the 3,456 tested cases were RDT-positive (Figure 1). Testing was not done when RDTs were stocked out, patients were admitted at nighttime and discharged by morning, patients refused, or the technician was ill. Percent positivity among those tested ranged from 36.6% to 46.9% (Table 1). Stable, seasonal transmission was observed with seasonal outbreaks typically beginning in the dry season (at the end of June/July to early October), followed by lower transmission in the rainy season (October—March/April) (Appendix Figures 5, 6). In some years, multiple peaks were seen in February and in the second half of the year between August and October (Figure 1). Peaks typically reached 80—100 weekly suspected cases. Earlier transmission starting in March was seen in 2019 and 2020.

**Table 1.** Description of testing among suspected cholera cases, Uvira, 2016 to 2020.

|  | 2016 | 2017 | 2018 | 2019 | 2020 |
| --- | --- | --- | --- | --- | --- |
| No. suspected cases | 1341 | 1134 | 1000 | 922 | 1050 |
| No. suspected cases per 10,000 population | 47.9 | 40.5 | 35.7 | 32.9 | 37.5 |
| No. (%) suspected cases that were RDT-tested | 617<br>(46.0) | 857<br>(75.6) | 533<br>(53.3) | 597<br>(64.8) | 852<br>(81.2) |
| No (%) RDT-positive cases among RDT-tested | 226<br>(36.6) | 374<br>(43.6) | 233<br>(43.7) | 260<br>(43.6) | 400<br>(46.9) |

Twenty-six spatiotemporal clusters were detected (Table 2). The mean cluster radius was 652m (range, 308—1582), mean size was 20 cases (range, 4—48), and mean duration was 24.8 days (range 1—58). The annual comparison of clustering demonstrated clustering in similar locations each year (Figure 2). The date of the first day of a retrospectively detected cluster usually (though not always) anticipated a surge in transmission over the next weeks, for all seasonal outbreaks except for early 2016 and 2017 when there were few cases tested (Figure 3, top). The median delay to the early warning signal (i.e., the number of days between retrospective detection date with all available data and the earliest prospective detection date) was 1 day (IQR 0—3) with a maximum delay of 23 days (Table 2). The median cluster size at signal detection was 3 cases (IQR 2—7) with a maximum size of 21 cases. Persistent clustering was seen in the north-central and southern areas, some distance away from the CTC (Figure 3, bottom). The 2016—2020 scan did not show clusters persisting between years but found larger clusters of 175—226 cases in the same locations. Sensitivity analyses of suspected cases found more clusters (N=32) of a similar mean radius and range (590m, range 270—1557) and larger mean size and range (37 cases, range 2—130) and longer duration (27.8 days, range 5—59), in similar locations (Appendix Table 2, Appendix Figure 7).

**Figure 2.**
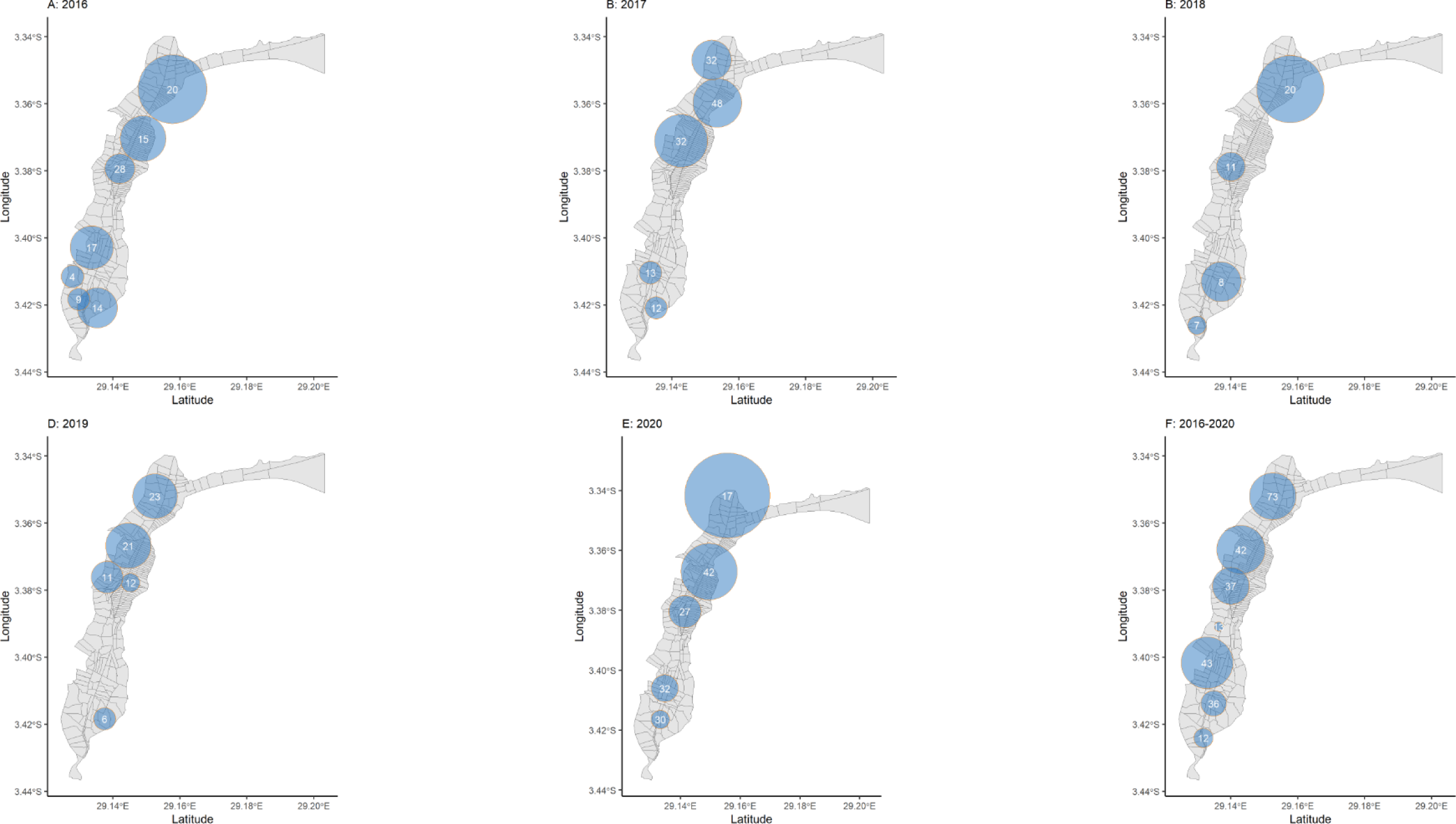
Spatial distribution of spatiotemporal clusters of RDT-positive cholera cases at the avenue level, Uvira, 2016— 2020. Clusters have a relative risk >1, p<0.05. The size of the orange circle depicts the spatial radius and the number of cases, in white.

**Figure 3.**
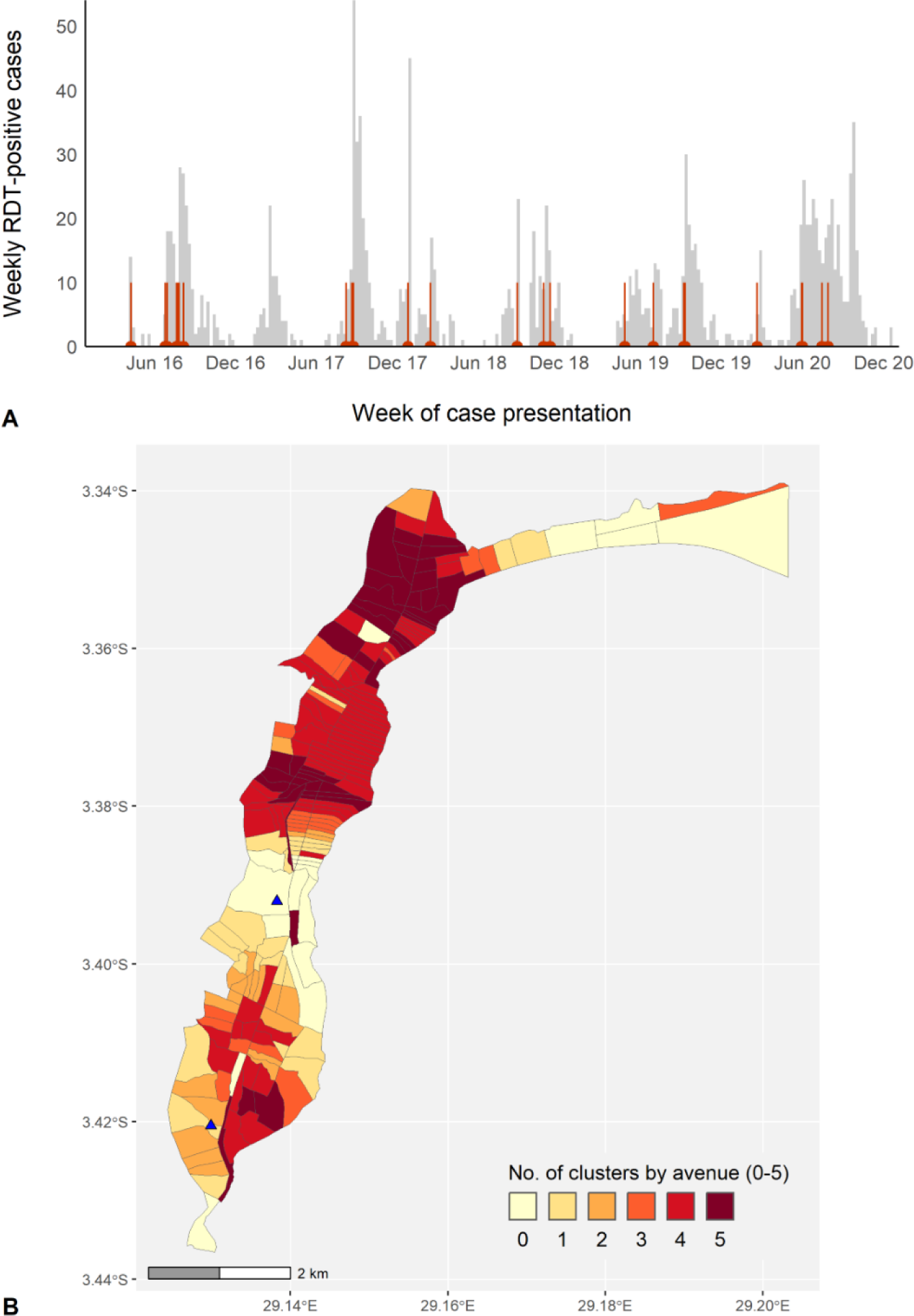
Cholera, Uvira, 2016—2020: (A) Epidemic curve showing weekly number of RDT-positive cholera cases based on week of presentation and start dates of 26 clusters (red vertical lines), (B) Cluster persistence within avenues for RDT-positive cases showing the number of years affected by clustering within avenues. The cholera treatment center (top) and unit (bottom) are marked with blue triangles.

**Table 2.** Spatiotemporal clusters of RDT-positive cholera cases detected through annual scanning at the avenue level, Uvira, 2016—2020.

| Year | No. | Cases observed: expected | Population at-risk | RR<sup>†</sup> | Cluster radius (m) | Cluster start date (mm/dd) | Cluster duration (days) | Signal delay (days)<sup>‡</sup> | Size at signal (cases) |
| --- | --- | --- | --- | --- | --- | --- | --- | --- | --- |
| 2016 | 1 | 20:1 | 30553 | 20.9 | 1140 | 08/05 | 18 | 8 | 11 |
|  | 2 | 28:3 | 34232 | 10.5 | 497 | 06/25 | 48 | 0 | 2 |
|  | 3 | 17:1 | 30758 | 13.8 | 717 | 07/22 | 23 | 5 | 12 |
|  | 4 | 15:1 | 31240 | 11.9 | 758 | 06/29 | 23 | 1 | 4 |
|  | 5 | 4:0 | 6579 | 344.4 | 376 | 04/09 | 1 | 0 | 3 |
|  | 6 | 14:2 | 30082 | 8.8 | 668 | 07/21 | 30 | 0 | 3 |
|  | 7 | 9:1 | 27452 | 12.6** | 368 | 07/26 | 14 | 3 | 4 |
| 2017 | 1 | 48:4 | 51012 | 13.0 | 811 | 08/07 | 40 | 2 | 2 |
|  | 2 | 32:2 | 43992 | 16.4 | 657 | 08/20 | 23 | 1 | 13 |
|  | 3 | 32:4 | 49794 | 7.7 | 880 | 08/23 | 44 | 0 | 2 |
|  | 4 | 13:1 | 51016 | 16.4 | 378 | 12/24 | 7 | 0 | 2 |
|  | 5 | 12:2 | 50635 | 7.6** | 368 | 08/23 | 15 | 12 | 2 |
| 2018 | 1 | 20:1 | 28884 | 26.6 | 1116 | 10/26 | 13 | 6 | 9 |
|  | 2 | 11:1 | 31204 | 22.7 | 475 | 02/13 | 7 | 0 | 3 |
|  | 3 | 8:0 | 25148 | 40.6 | 662 | 08/28 | 3 | 0 | 4 |
|  | 4 | 7:0 | 17345 | 18.6** | 308 | 11/10 | 10 | 1 | 3 |
| 2019 | 1 | 23:1 | 33751 | 18.6 | 743 | 09/10 | 18 | 1 | 7 |
|  | 2 | 21:3 | 33162 | 9.0 | 755 | 09/07 | 35 | 0 | 12 |
|  | 3 | 12:1 | 16210 | 12.3 | 309 | 04/27 | 29 | 1 | 2 |
|  | 4 | 11:1 | 16495 | 13.2 | 527 | 09/07 | 24 | 0 | 2 |
|  | 5 | 6:0 | 15001 | 27.8** | 368 | 06/30 | 6 | 0 | 2 |
| 2020 | 1 | 42:6 | 60378 | 7.8 | 1048 | 07/29 | 58 | 2 | 3 |
|  | 2 | 27:3 | 42423 | 8.7 | 599 | 07/15 | 46 | 23 | 21 |
|  | 3 | 17:1 | 56029 | 19.1 | 1582 | 02/20 | 9 | 0 | 2 |
|  | 4 | 30:5 | 63207 | 6.5 | 343 | 05/30 | 46 | 2 | 6 |
|  | 5 | 32:6 | 63593 | 5.8 | 501 | 06/01 | 55 | 4 | 6 |

\* p-value < 0.001 ≥ p-value < 0.01; \*\* p-value < 0.001. <sup>†</sup> RR, relative risk. <sup>‡</sup> Signal delay indicates the number of days between retrospective detection date with all available data and the earliest prospective detection date.

Among RDT-positive cases from 2016—2020, within the first 5 days after a case presented for care, the high-risk zone extended from the case residence to 1105m and the risk remained elevated up to 1665m (maximum moving average τ = 1.8, 95% CI 1.4—2.3, Figure 4A). During days 1—4, when a response can be more realistically mobilized, the risk zones remained similar (Figure 4B). Examining RDT-positive cases in 2020 alone, the high-risk zone extended to a smaller radius of 585m, and the risk remained elevated up to 1915m (maximum moving average τ = 1.8, 95% CI 1.0—2.9, noting the wider confidence intervals for the smaller 2020 dataset, Figure 4C). During days 1—4, the risk zones were 425m and 1915m, respectively (maximum moving average τ = 1.7, 95% CI 1.1—2.6, Figure 4D). In the sensitivity analysis of suspected cases from 2020, the trends remained like RDT-positive cases in 2020 (Figure 4C, 4D versus 4E, 4F). For suspected cases, during days 1—4, these risk zones were 1155m and 2075m, respectively (maximum moving average τ = 1.6, 95% CI 1.3—2.0, though a drop in the lower CI was observed at 635m, as marked by first vertical dashed line in Figure 4F). Results by year for RDT-positive cases showed lower ranges in the radii of the high-risk zone (425m across all years except 2017 where it is 875m) and the elevated zone (1125—1485m) (Appendix Figure 8). Using simulated individual household locations (from 75—2500m for days 0—4), the results were similar to the main analysis of centroid locations with a moving average τ ≥ 2.0 measured from 75m to 275m (maximum moving average τ = 2.4, 95% CI 1.7—3.3), a similar high-risk zone radius of 1415m, and a similar descending trend in risk over distance, central tendencies and correlation coefficients (Appendix Figure 4, Appendix Table 1).

**Figure 4.**
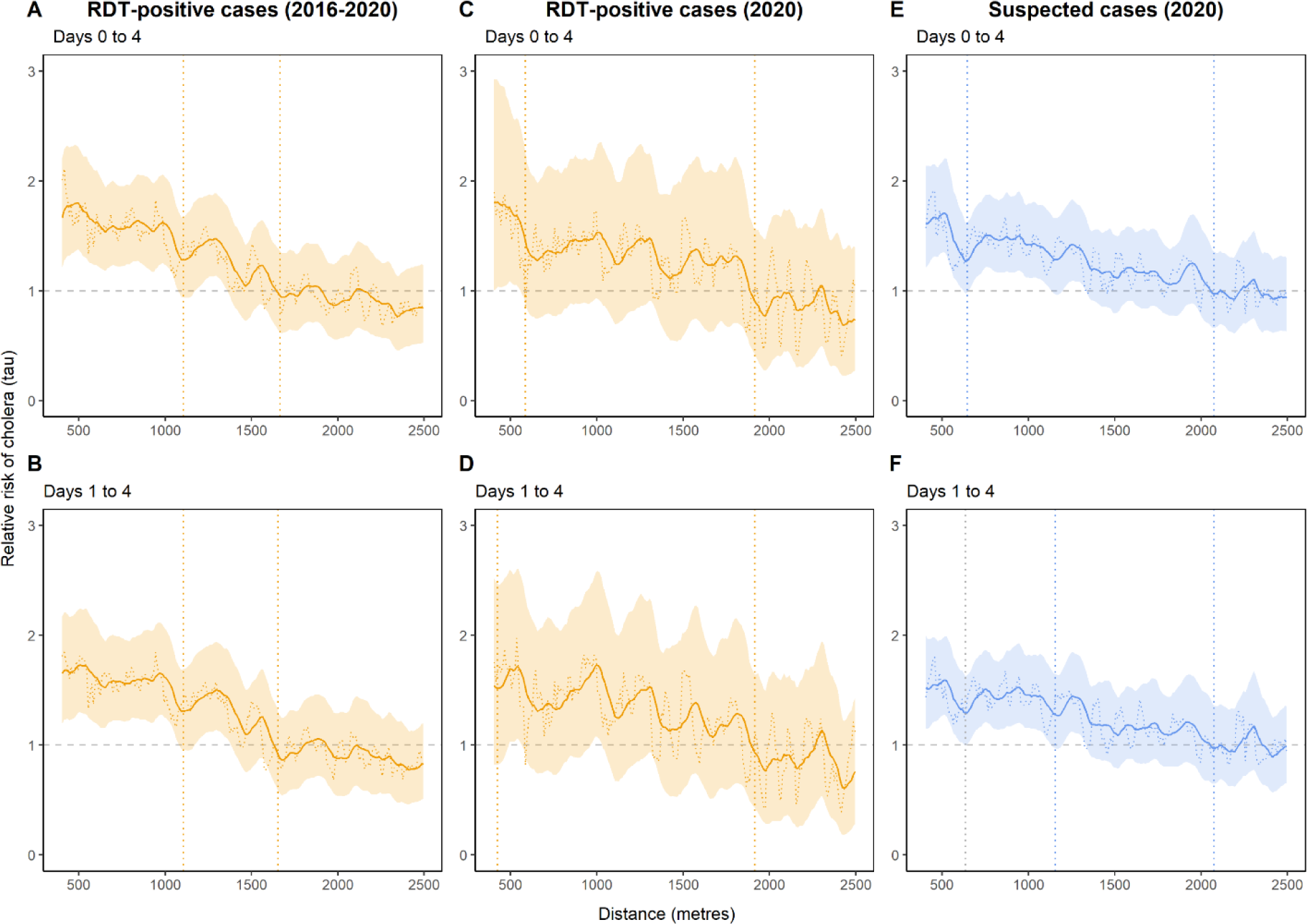
Cholera, Uvira, 2016—2020: Moving average estimates of τ (relative risk) and 95% CIs (solid line and shading) with point estimates (dashed horizontal line) for days 0—4 (panels A, C, E) and days 1—4 (panels, B, D, F), for RDT-positive cases (in orange) and suspected cases (in blue) for cholera in 2016—2020 (panels A, B) and (panels C, D, E, F), using 1000 bootstrap samples. The vertical dashed lines indicate the spatial extent of the zone of high-risk where the lower 95% CI crossed 1.0 for ≥30m consecutively (first line) and zone of elevated risk where the point estimate crossed 1.0 for ≥30m consecutively (second line).

## DISCUSSION

We provide insight into clustering dynamics from one of the world’s most burdensome cholera hotspots. Elucidating clustering patterns in an endemic setting can specify where intensive transmission is occurring early on, within small areas. Our results suggest that targeted interventions can take advantage of this natural clustering to mitigate seasonal outbreaks before they enlarge. Cities like Uvira, Goma and Bukavu are thought to regularly seed regional outbreaks; investigating transmission routes and coordinating prevention and control strategies there can have substantial public health benefits.(18)

The two clustering methods produced aligned results. When evaluating the 2016—2020 period, a 1105m high-risk radius around RDT-positive cases within the 5 days after case presentation was estimated with a τ ≤ 1.8. The risk zone for 2020 showed a 600m high-risk radius with a τ ≤ 1.8. These radii are consistent with those estimated for Matlab, Bangladesh (500m, within 4—6 days post-presentation, RR = 1.9).(10) In Uvira, an elevated-risk radius up to 2000m around cases demonstrates the persistence of risk in a densely-populated city. The risk zones remained intact after a 1- day delay wherein it is realistic to launch a response. The τ estimates are higher than estimates from N’Djamena (220m) and Kalemie (330m), likely reflecting propagation among neighboring households, but it should be noted that we do not include distances <420m.(9) To explore this omission further, the simulation of household locations produced similar risk zones and an initial increase in risk from 75—275m equivalent to τ < 2.5. There may be additional epidemiological differences including increased environmentally-mediated transmission, immunity, population density, and mobility related to seasonal fishing and trading.

The space-time scan statistic demonstrated a mean radius (650m) among the 26 spatiotemporal clusters which emerged in the south and central-north areas. This is similar to the τ high-risk zone for 2020 (600m). The start date of the retrospectively- detected cluster acted as an alarm that usually preceded the onset of seasonal outbreaks. The early warning signal for these clusters was delayed by a median of 1 day (whereas delays of >1 week for 3 clusters are not feasible for rapid response) Our study’s main strength is the use of high-specificity RDT-positive cases as compared to suspected cases alone. Previous analyses have relied on suspected cases which may overestimate risk due to inclusion of other diarrheal pathogens.(9–11) Our sensitivity analyses based on suspected cases showed similar results but with likely false positive clusters detected and a larger radius. In Uvira, the CTC/CTU may not be the main source of care for diarrhea, particularly when mild. In a 2021 community survey of health-seeking practices among Uvira residents, most persons with any diarrhea in the past week (70%) reported that they first visited pharmacies for care, rather than CTCs (4%).(41) Use of medically-attended cases introduces a potential bias of including only moderate to severely-ill cases, and therefore missing the transmission from milder cases. This is mitigated if medically attended cases represent a random proportion of all cholera cases. A major limitation is that the spatial resolution is based on avenue centroids, not household locations. This misses household transmission and case-pair distances <420m, where 5% of distances fell. Our simulations of household locations showed qualitatively similar trends, with higher τ at a smaller radius. Other limitations of the τ statistic include limited power to detect true risk areas using narrow distance bands where the sample size of related pairs is small.(42) The τ trendline and its sampling error are not smooth, as the clustering algorithm is recalculated every 50m and the moving average recalculated every 10m. Given that annual estimates are based on <500 RDT-positive cases, evaluation of the minimum number of cases needed for reliable τ estimation is needed.(42) For both τ and the space-time scan statistic, a circular radius has reduced sensitivity to detect the true geographical extent of noncircular clustering or an outbreak (i.e., an outbreak along the coastline, as might be the case in Uvira given its lakeside position), though detection appears unaffected.(34)

The results can inform control measures for seasonal outbreaks. The mapping of persistent clustering can be used to prioritize persistent high-recurrence hotspots for preventative measures, where transmission occurs early and predictably. Aiming for high coverage and uptake of preventative interventions in these areas can reduce exposure, reinfection, and transmission. Daily prospective scanning for local clustering could aid in early cluster detection across Uvira.(38) The radii of 100—500m used for a CATI strategies in DRC(17, 43) are justified by these findings and could perhaps be enlarged further. A 600—1105m radius of infection risk would include several thousand persons and would be logistically prohibitive to cover rapidly (i.e., within the 3 to 5 day risk window). However, CATIs may be considered for early containment of (a) potential zones of infection around new cases in less affected areas that fall outside of the known high-recurrence areas, (b) small outbreaks among lakeside communities in hotspots which may seed larger outbreaks(18), and (c) sporadic cases after mass vaccination(15). Building on previous and current studies(9, 10) and operational experience(17, 43), a 200—600m radius can be used to narrow down areas for CATIs where transmission is likely.

## Acknowledgements

We thank the Uvira Health Zone and CTC/CTU collaborators for the support provided to testing and data collection, often under difficult circumstances. We thank John Giles, University of Washington, for advice on implementing the IDSpatialStats package. Last but not least, we thank all the patients who agreed to participate in the main trial.

## Data availability

All data and code produced are available online at https://github.com/ruwanepi/Uvira_spatiotemporal

