## Appendix for "Spatiotemporal modelling of cholera and implications for its control, Uvira, Democratic Republic of the Congo"

### Appendix Text 1: Statistical framework for local and global clustering statistics

#### Methods for the space-time scan statistic(1, 2)

For a given cylinder consisting of a radius centered on an avenue-centroid and height of the temporal window of interest, ***c*** is the observed number of cases inside the cylinder, ***E[c]*** is the expected number of cases for any given cylinder, and ***C*** is the total number of cases in Uvira, with $RR$ given by:

$$RR = \frac{\frac{c}{E[c]}}{\frac{(C - c)}{(C - E[c])}}$$

During the scan, a circular scanning window of varying radii and duration moves over the geographical area, so that each avenue-centroid is at the center of several candidate clusters of differing radii and heights. At each cylinder location, the number of cases inside the cylinder is compared with the expected number, under a null hypothesis of no clustering (i.e., cases are randomly distributed). To find the most likely cluster, candidate clusters are ordered by a log-likelihood ratio (LLR) where the cluster with the largest LLR is the least likely to be due to chance and therefore, the most likely cluster. The significance of each cluster was evaluated using Monte Carlo simulation to compare the original dataset with 999 random replicates produced under the null hypothesis.

**Methods for the $\tau$ statistic(3-5)**

$\hat{\tau}(d_1, d_2)$ as an RR is approximated by dividing the odds that cases within the band are transmission-related $\hat{\theta}(d_1, d_2)$ by the same odds among cases in the general population, regardless of distance $\hat{\theta}(0, \infty)$.

The $\tau$ equation is given by: $\hat{\tau}(d_1, d_2) = \frac{\hat{\theta}(d_1, d_2)}{\hat{\theta}(0, \infty)}$

The odds for numerator $\hat{\theta}(d_1, d_2)$ are given by: $\hat{\theta}(d_1, d_2) = \frac{\sum i \sum j * I_1(i,j)}{\sum i \sum j * I_2(i,j)}$

The numerator tallies the number of case-pairs (i-j) within the given distance band that are transmission-related (within 0—4 days) (using indicator variable $I_1(i, j)$=1 for notation). The denominator tallies the number of case-pairs (i-j) within the given distance band that are not transmission-related (occurring after 4 days) (using indicator variable $I_2(i, j)$=1 for notation). The equivalent odds $\hat{\theta}(0, \infty)$ is estimated for the entire population.

### Appendix Text 2: Simulations to compare centroid-geotagged cases with cases with simulated individual household locations

The case data used in this study are geocoded by X/Y coordinates of the centroid of the 216 avenues (or streets) of the case's residence (Appendix Figure 1 displays the avenue boundaries and their centroids). In this simulation, we assess whether using centroids versus simulated individual household locations affects trends in the tau statistic, and to what extent.

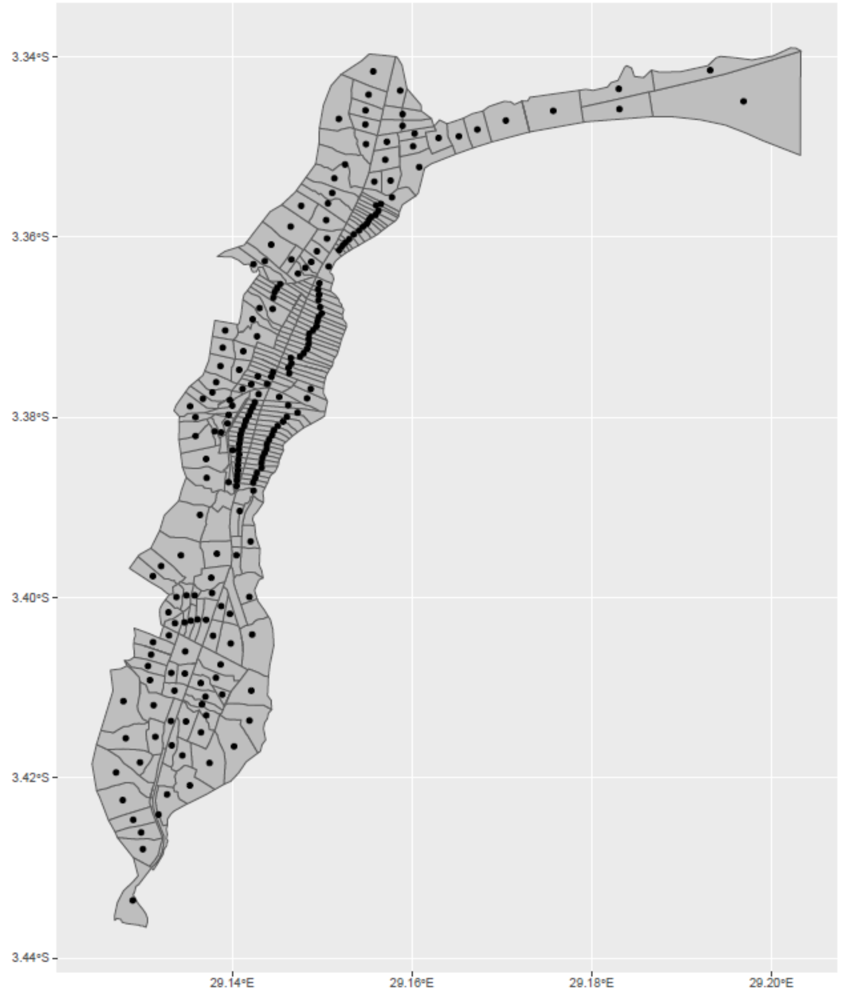


**Appendix Figure 1.** Map of the centroid locations and borders of Uvira's 216 avenues

### Methods

We used the dataset of 1493 rapid diagnostic test (RDT) positive cholera cases from 2016—2020 (displayed in space and time in Appendix Figure 2). The X/Y coordinates in this dataset were perturbed randomly by adding a random normal distribution with an arbitrarily-defined standard deviation of 100. The points were plotted as maps to visually compare the spatial spread of cases between datasets 1 and 2 (Appendix Figure 3). The main $\tau$ analysis was run for each dataset. This produced the $\tau$ statistic (relative risk and 95% CIs) of the next case being within a specific distance to another case (y-axis) compared with the risk of the case occurring anywhere else during days 0—4 for RDT-positive cases. A moving average was applied in distance spans of 10m, 25m, and 50m to smooth fluctuations. To assess the similarity between the datasets, the trendlines were evaluated visually by graphing and by comparing Pearson correlations.

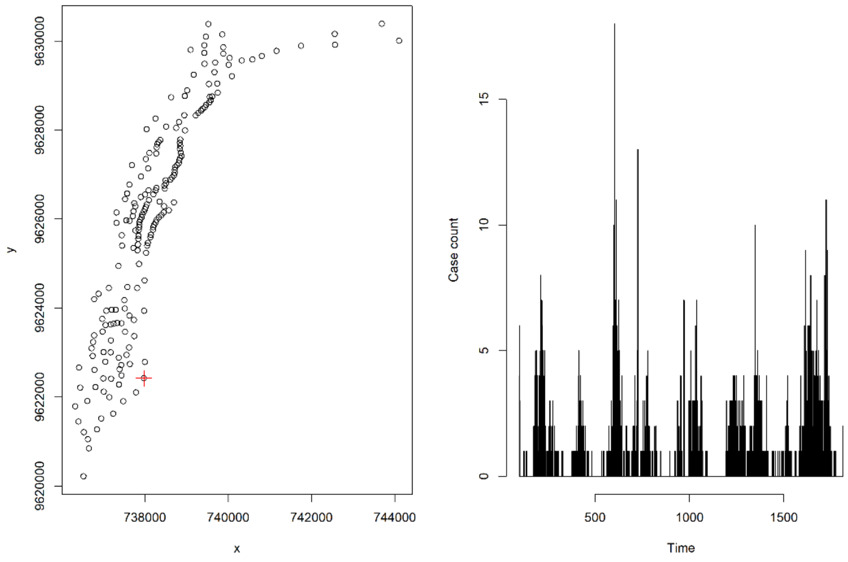


**Appendix Figure 2.** Uvira 2016—2020 dataset of rapid diagnostic positive cases with avenue centroids of cases (index case in red)

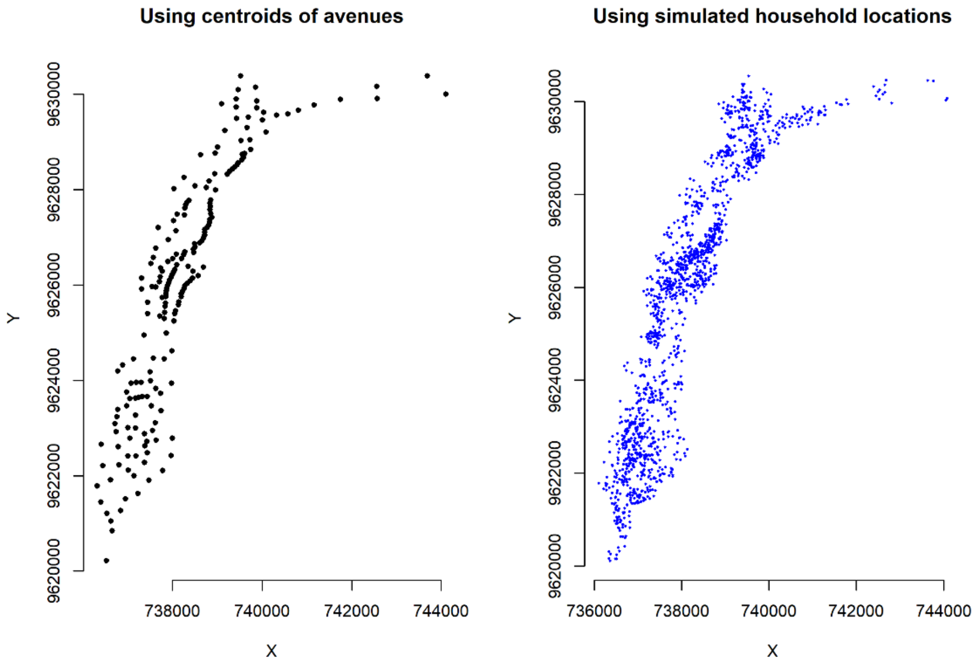


**Appendix Figure 3.** Case centroid locations (black) and simulated household locations (blue)

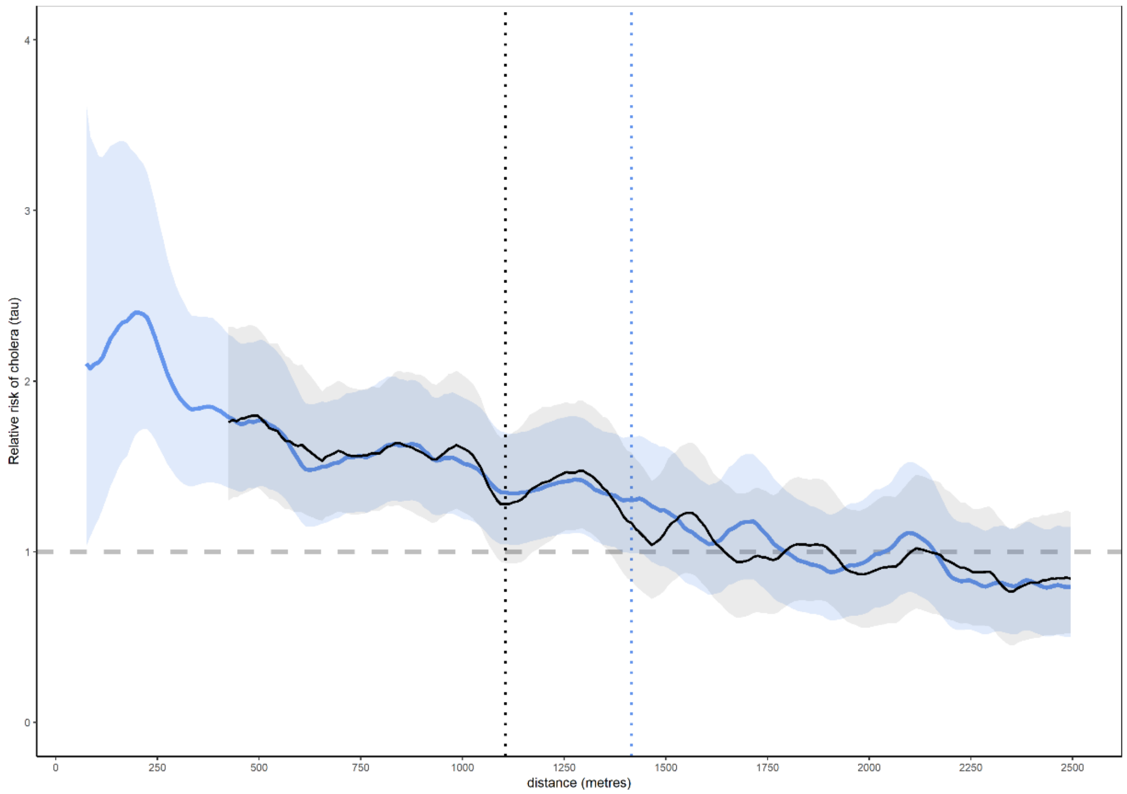


**Appendix Figure 4.** Moving average of point estimates and 95% confidence intervals for tau $\tau$ statistic for RDT-positive cholera cases (75—2500m) of the centroids (black, starting at 420m) and the household locations (blue, starting at 75 m). The dashed line is where the lower confidence interval for the moving average crosses 1.0 three times consecutively.

### Findings and interpretation

The two datasets showed similar $\tau$ trends (Appendix Figure 4). Both for lower CIs of the moving average $\tau$ and for the moving average $\tau$ point estimates where $\tau$ crossed 1.0 twice consecutively differed between the centroid dataset and the household dataset (Appendix Table 1). The Pearson correlation coefficients were significant and nearly identical.

**Appendix Table 1.** Differences in points where $\tau$ crosses RR=1.0 twice consecutively

| Dataset | Min τ | Max τ | Mean τ | Moving average τ <1.0 (3 times) | Moving average τ LCI <1.0 (3 times) | Pearson correlation coefficient |
|---|---|---|---|---|---|---|
| Centroid | 0.52 | 3.01 | 1.01 | 1665m | 1105m | -0.87 (95% CI -0.89, -0.85) |
| Simulated household | 0.55 | 2.40 | 1.05 | 1815m | 1415m | -0.88 (95% CI -0.90, -0.86) |

Overall, the centroid dataset showed a similar descending trend in risk over distance, central tendencies and correlation coefficients, as compared with the simulated household dataset. The centroid dataset however showed a lower $\tau$ threshold estimate for the moving average $\tau$ point estimate (by 8.3%) and lower 95% CI moving average (by 21.9%). Notably, the simulated households had the highest moving average $\tau$ estimate (equivalent to 2.0 < RR < 2.5) from 75—275m, which was unmeasured in the centroid dataset.

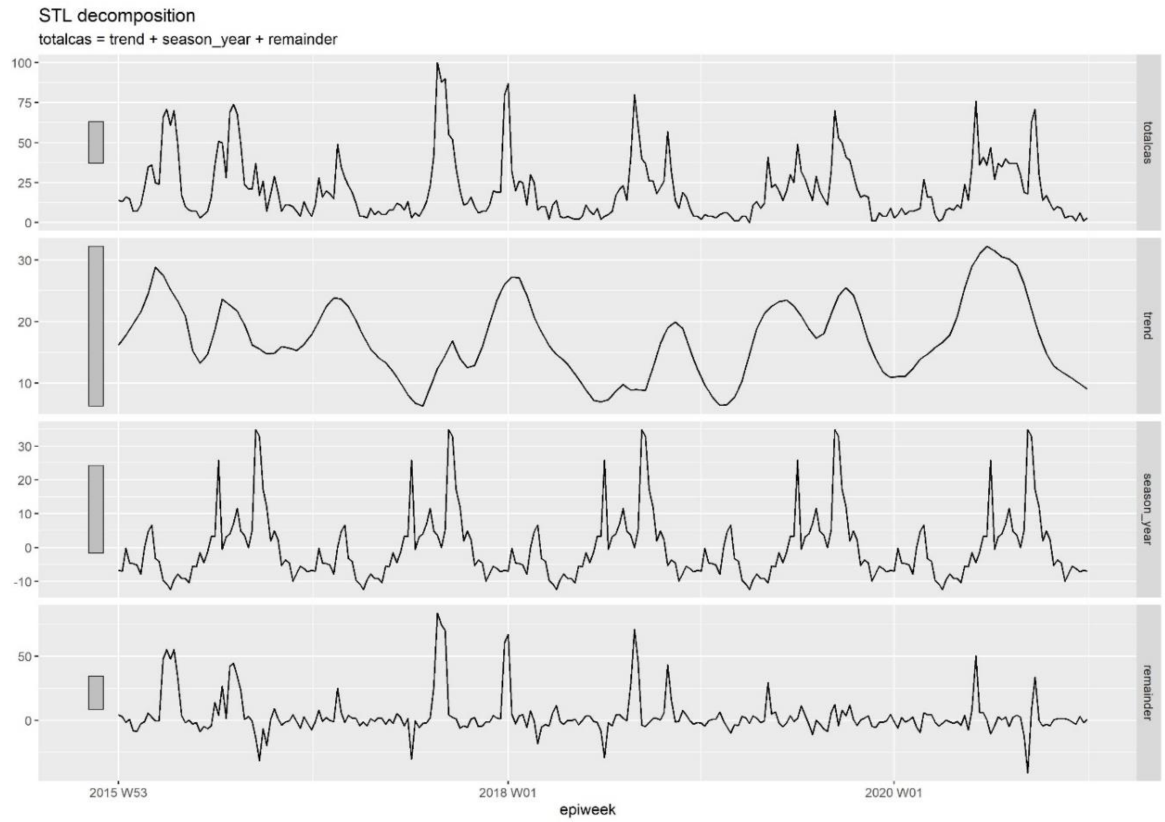


**Appendix Figure 5.** Trend, season and remainder decomposition using a trend window for smoothing of 14 days and seasonal window for smoothing including the entire period

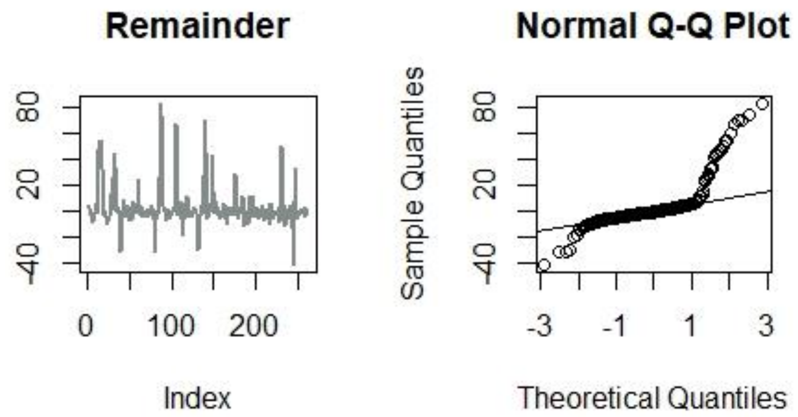


**Appendix Figure 6.** Normal Q-Q plot of residuals (remainder) and verification of a heavy-tailed skew approaching a normal distribution of residuals (indicating a mix of structure and noise)

**Appendix Table 2: Sensitivity analysis: spatiotemporal clusters of suspected cholera cases, Uvira, 2016—2020**

| Year | No. | Cases observed | Cases expected | Population at-risk | RR | Radius (m) | Start date (mm/dd) | Duration (days) |
|---|---|---|---|---|---|---|---|---|
| 2016 | 1 | 57 | 5 | 177122 | 10.8 | 378 | 04/07 | 15 |
| | 2 | 51 | 4 | 187076 | 12.1 | 647 | 03/24 | 11 |
| | 3 | 45 | 6 | 183225 | 7.2 | 1557 | 08/06 | 17 |
| | 4 | 27 | 3 | 120498 | 8.4 | 368 | 04/09 | 13 |
| | 5 | 40 | 9 | 147424 | 4.6 | 709 | 07/22 | 30 |
| | 6 | 18 | 2 | 29390 | 7.8 | 436 | 02/18 | 40 |
| 2017 | 1 | 130 | 13 | 148014 | 10.8 | 908 | 08/07 | 43 |
| | 2 | 91 | 16 | 150104 | 5.9 | 897 | 08/19 | 52 |
| | 3 | 39 | 6 | 88959 | 6.6 | 704 | 08/29 | 32 |
| | 4 | 23 | 2 | 134147 | 10.6 | 378 | 12/24 | 7 |
| | 5 | 26 | 5 | 143948 | 5.2 | 1001 | 08/23 | 16 |
| | 6 | 9 | 1 | 42275 | 17.3 | 331 | 02/14 | 5 |
| 2018 | 1 | 50 | 3 | 130673 | 15.3 | 963 | 10/26 | 12 |
| | 2 | 24 | 2 | 134311 | 15.1 | 397 | 01/01 | 5 |
| | 3 | 61 | 15 | 132515 | 4.2 | 906 | 07/29 | 56 |
| | 4 | 44 | 10 | 128631 | 4.5 | 708 | 08/21 | 38 |
| | 5 | 18 | 3 | 70142 | 5.9 | 653 | 10/30 | 21 |
| | 6 | 9 | 1 | 52203 | 14.4 | 477 | 02/17 | 5 |
| 2019 | 1 | 50 | 4 | 93453 | 14.3 | 831 | 09/10 | 18 |
| | 2 | 30 | 2 | 21965 | 13.9 | 0 | 09/01 | 48 |
| | 3 | 47 | 7 | 105035 | 7.1 | 524 | 04/27 | 31 |
| | 4 | 48 | 10 | 115699 | 5.0 | 836 | 09/07 | 41 |
| | 5 | 36 | 8 | 120197 | 4.7 | 995 | 06/08 | 31 |
| | 6 | 14 | 2 | 40341 | 7.4** | 626 | 06/23 | 22 |
| | 7 | 6 | 0 | 45292 | 32.2 | 350 | 09/20 | 1 |
| 2020 | 1 | 105 | 17 | 159204 | 6.7 | 860 | 07/29 | 59 |
| | 2 | 59 | 11 | 141671 | 5.8 | 488 | 05/31 | 41 |
| | 3 | 38 | 5 | 106256 | 8.6 | 1121 | 02/20 | 23 |
| | 4 | 57 | 13 | 155765 | 4.6 | 395 | 05/30 | 46 |
| | 5 | 49 | 13 | 120618 | 3.9 | 490 | 07/27 | 59 |
| | 6 | 39 | 10 | 159261 | 4.0 | 959 | 05/30 | 34 |
| | 7 | 15 | 2 | 44366 | 10.1 | 468 | 09/10 | 18 |

* p-value < 0.001 ≥ p-value <0.01; ** p-value < 0.001. † RR, relative risk. ‡ Signal delay indicates the number of days between retrospective detection date with all available data and the earliest prospective detection date.

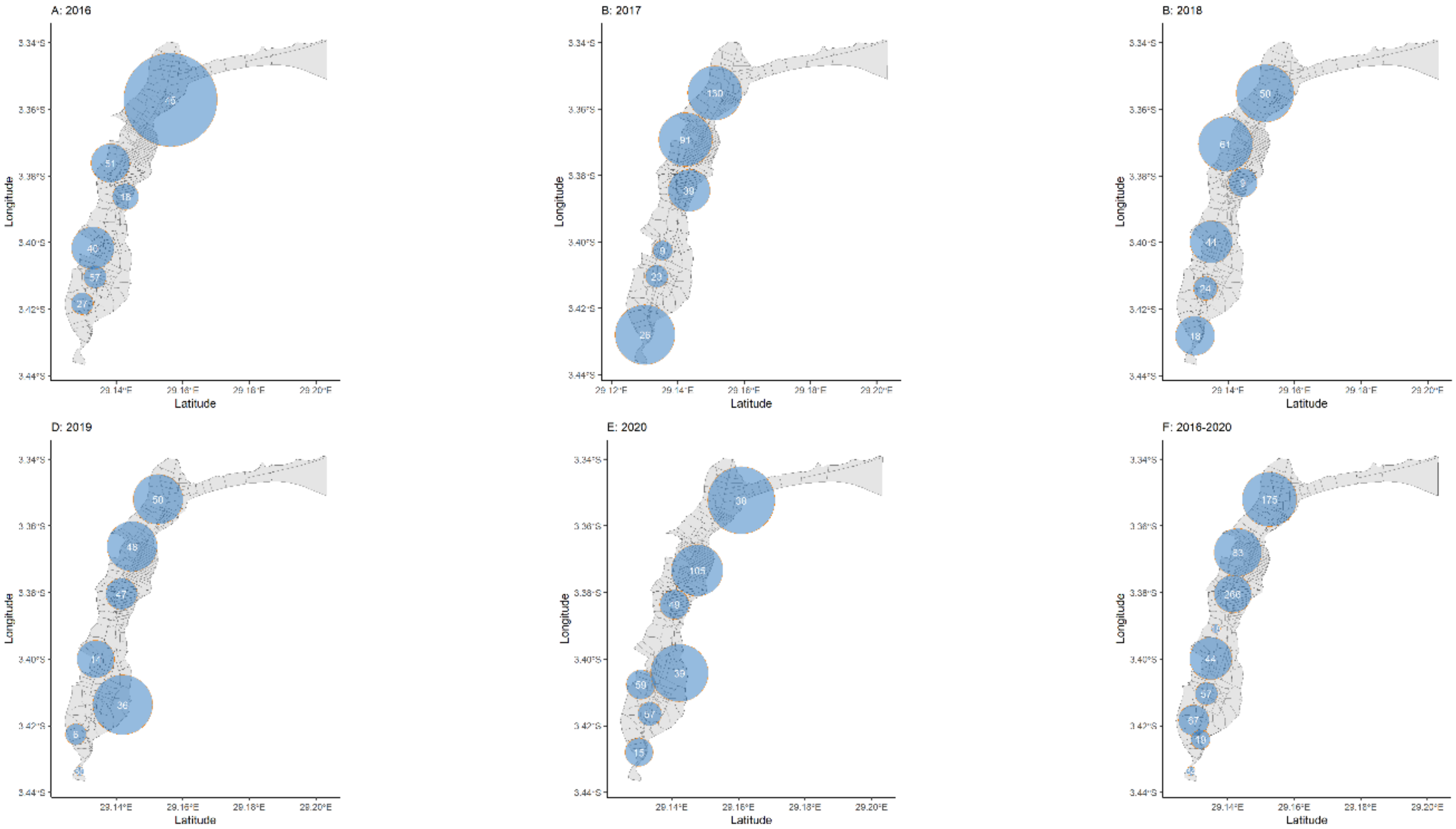


**Appendix Figure 7.** Sensitivity analyses of prospectively detected spatiotemporal clusters of suspected cholera cases, 2016—2020. The size of the orange circle depicts the radius with the number of suspected cases (in white). A—E depict scans with a temporal window of 7—60 days and F depicts a scan with a temporal window of 7—365 days. All scans had a maximum spatial window of 10% of the geographical area.

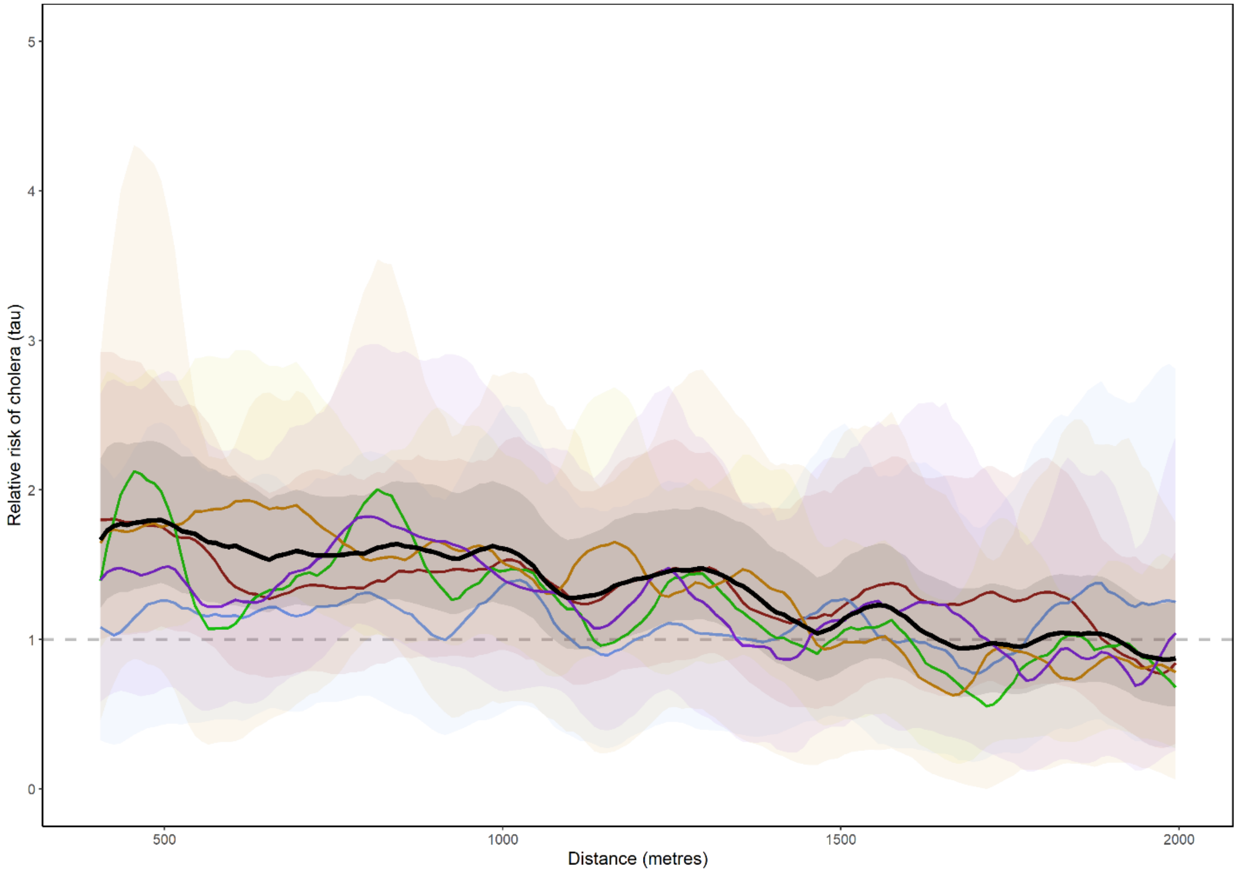


**Appendix Figure 8.** Cholera, Uvira, 2016—2020: Annual and aggregated moving average estimates of $\tau$ (relative risk) and 95% CIs (solid line and shading) for days 0—4. 2016—2020 in black, 2016 in purple, 2017 in orange, 2018 in green, 2019 in blue, 2020 in red.
